# Results of a phase Ib study of SB-121, an investigational probiotic formulation, in participants with autism spectrum disorder

**DOI:** 10.1101/2022.11.14.22281882

**Authors:** Lauren M. Schmitt, Elizabeth G. Smith, Ernest V. Pedapati, Paul Horn, Meredith Will, Martine Lamy, Lillian Barber, Joe Trebley, Kevin Meyer, Mark Heiman, Korbin H.J. West, Phoevos Hughes, Sanjeev Ahuja, Craig A. Erickson

## Abstract

Autism Spectrum Disorder (ASD) is a neurodevelopmental disorder characterized by core impairments in social communication as well as restricted, repetitive patterns of behavior and/or interests. Individuals with ASD, which includes about 2% of the US population, have challenges with activities of daily living and suffer from comorbid medical and mental health concerns. While aripiprazole and risperidone are indicated for use in autistic youth with interfering irritability, there are no drugs indicated for the core impairments of ASD. As such, there is a significant need for the development of new medication strategies for individuals with ASD. This first-in-human placebo-controlled, double-blind, crossover study investigated the safety (primary objective) and efficacy of oral SB-121, a combination of *L. reuteri*, Sephadex^®^ (dextran microparticles), and maltose administered once daily for 28 days in 15 autistic participants. SB-121 was safe and well tolerated. SB-121-associated directional improvements in adaptive behavior measured by Vineland-3 and social preference as measured with eye tracking were noted. These results provide support for further clinical evaluation of SB-121 as a treatment in autistic patients.

---

Autism Spectrum Disorder (ASD) is a neurodevelopmental disorder characterized by core impairments in social communication and interaction combined with restricted, repetitive patterns of behavior and/or interests [1]. It is estimated that 1.7-2.2% of people of all ages are diagnosed with ASD in the United States [2,3]. Individuals with autism may struggle to function at school, work, and in everyday life situations. These challenges may be compounded by high rates of comorbid mental and physical health conditions [4-7]. These include, but are not limited to, gastrointestinal (GI), immunological, and psychiatric disorders. Overall, autistic individuals are at a 3 to 10 times higher risk for premature mortality [8,9] compared to the general population. These issues highlight the critical need for the development of treatment options in ASD [10].

Despite decades of research focused on the development of therapeutics for the treatment of the core social, communication, or functional impairments associated with autism, no such drugs have been approved by the United States Food and Drug Administration (FDA) [11]. The only approved drugs in ASD are aripiprazole and risperidone, which are limited to the treatment of irritability associated with physical aggression, self-injurious behavior, and severe tantrums in autistic youth [12].

Enhancement of oxytocin signaling is among the more promising and well-studied targets of core symptom treatment development in ASD[11]. Oxytocin is an endogenous neuroendocrine hormone produced in the hypothalamus, released by the posterior pituitary into blood and stimulates milk letdown and uterine contractions in females. Hypothalamic oxytocin neurons also project to areas within the central nervous system (CNS) that are responsible for regulating social behavior[13-15]. Central oxytocin pathways also may project to the efferent vagal nervous system. In rats, intracisternal oxytocin administration reduces colonic hyperpermeability via the vagal cholinergic pathway[16]. The relationship between oxytocin activity and gastrointestinal symptomatology has not been directly explored in ASD.

Several studies have specifically investigated the oxytocin system in autism. A meta-analysis of studies using plasma oxytocin as a biomarker in ASD noted that many, but not all, autistic youth showed reduced oxytocin levels[17]. A meta-analysis of oxytocin receptor gene single-nucleotide polymorphisms noted an association between autism and certain polymorphisms[18]. However, exogenously administered oxytocin does not cross the blood-brain barrier. While this may potentially be overcome by the intranasal administration of oxytocin, clinical trials of intranasal administration of oxytocin in ASD have demonstrated mixed results [11]. Improvements in emotion recognition and social behavior were noted in several early phase trials [19-22], while a large 24-week double-blind placebo-controlled parallel group trial of intranasal oxytocin in 290 autistic youth noted no treatment-associated positive clinical effects [23]. Study authors hypothesized that intranasal oxytocin administration may not adequately mimic the endogenous pulsatile oxytocin pattern and stimulation of the CNS oxytocin receptor, which may have contributed to lack of efficacy in this trial [24,25]. Given the importance of oxytocin in regulating social behavior and the challenges with exogenous administration, there is a clear need to evaluate alternative approaches to enhancing endogenous neuronal secretion of oxytocin in autistic individuals.

Studies show a high prevalence of gastrointestinal symptoms in patients with ASD, with autistic youth almost eight times more likely to suffer from significant gastrointestinal symptoms such as constipation, GI pain, or diarrhea than those with typical development [26]. Additionally, gastrointestinal symptoms in autistic patients have been demonstrated to correlate with the degree of maladaptive behavior such as irritability, social withdrawal, hyperactivity, and interfering repetitive behavior [26].

*Limosilactobacillus reuteri (L. reuteri)* [Lr], formerly known as *Lactobacillus reuteri*, is a probiotic bacterium that naturally colonizes the outer mucous layer of the intestines. *L. reuteri* stimulates production of mucin by goblet cells and protects intestinal cells from opportunistic pathogens. Oral *L. reuteri* treatment has been associated with reduction in social deficits in three mouse models of ASD and is mediated via CNS oxytocin signaling [27]. It is hypothesized that these improvements have been driven by the ability of *L. reuteri* to stimulate oxytocin signaling to the ventral tegmental area of the CNS, a region with a significant role in reward, motivation, cognition, and aversion [27-30].

SB-121 is a formulation of *L. reuteri* with Sephadex^®^ (dextran microparticles, [DM]) and maltose. This combination results in a series of beneficial changes in the bacterium, including increased adherence to intestinal epithelial cells, improved gastric survival, and enhanced persistence through biofilm formation [31]. In this activated state, *L. reuteri* use has been associated with reduced disease incidence and severity, reduced intestinal inflammation and permeability, and reduced mortality in animal models of necrotizing enterocolitis (NEC) or *Clostridioides difficile* infection [32-34].

Given the need to evaluate therapeutic methods to safely boost endogenous CNS oxytocin signaling in patients with ASD, combined with the clear high rates of gastrointestinal dysregulation in individuals with autism, we proposed to evaluate the safety of oral administration of SB-121 in adolescents and adults with autism. Secondarily, we proposed to evaluate the potential efficacy of SB-121 in ASD.

## Methods

We conducted a randomized double-blind, placebo-controlled crossover trial of SB-121 in fifteen 15-45 year old autistic participants. This study was conducted at the Cincinnati Children’s Hospital under ICH GCP, and all applicable regulatory requirements. The protocol was reviewed and approved by the Cincinnati Children’s Hospital Institutional Review Board and registered at clinicaltrials.gov (NCT04944901). All participants under age 18 or over 18 with a legal guardian had a parent or legally authorized caregiver provided informed consent for their participation. Each participant provided their own additional consent or assent as possible and applicable. All the inclusion and exclusion criteria are listed in Supplementary Information. In short, all participants had a confirmation of DSM-5 criteria-based ASD diagnosis using the Autism Diagnostic Observation Schedule, 2^nd^ Edition (ADOS-2)[35]. Participants had to be free from active, uncontrolled GI symptoms or fever, and autoimmune disorders. Additional exclusion criteria included use of proton pump inhibitors, antibiotics, monoclonal antibodies, immunosuppressive drugs, and probiotics excluding yogurt. SB-121 (or placebo) was given daily and each dose consisted of 2 X 10^10^ colony forming units of *L. reuteri*, 200 mg of Sephadex^®^, and 74 mM of maltose in a final volume of 10.8 mL. Placebo consisted of 200 mg Sephadex^®^ and 74 mM of maltose in a final volume of 10.8 mL. After reconstitution of either SB-121 or placebo, the mixture was left for 15-45 minutes at room temperature and then consumed mixed with a preferred drink (water or juice).

Following consent, a screening visit was completed including administration of the ADOS-2 (which was not required if assessment was completed within the previous 36 months and results were available), clinical interview using DSM-5 criteria for ASD, a medical and psychiatric history, physical examination and laboratory tests done to confirm study eligibility. Following randomization, but before the initial dose of SB-121 or placebo, additional subject characterization measures were completed including cognitive testing using the Wechsler Abbreviated Scale of Intelligence, 2^nd^ Edition (WASI-II) and administration of the Social Communication Questionnaire (SCQ). Participants completed additional assessments both pre-dose and following 28 days of daily SB-121/placebo dosing (i.e., at outcome) including the Vineland Adaptive Behavior Scales, 3^rd^ edition (Vineland-3)[36], Aberrant Behavior Checklist (ABC)[37], and Clinical Global Impressions Severity (CGI-S) and Improvement (CGI-I; done post-treatment only) subscales[38]. Additionally, we evaluated quantitative subject performance using a social versus non-social scene preference eye tracking task[39]. Last, blood samples were collected for the evaluation of plasma oxytocin at baseline and following 28 days of treatment with SB-121 and placebo (i.e., at the start and end of each period for four total collections). A two-week washout period occurred between each treatment period.

Regarding safety evaluations, participants completed safety laboratory panels (hematology and blood chemistry studies) and vital signs pre- and post-28 days of treatment during each treatment period. A full physical examination was done at screening and subsequently a limited focused physical examination was done for the evaluation of adverse events, as needed at all in person visits. Adverse events, concomitant medications and treatment compliance were assessed during all visits. All treatment emergent adverse effects (TEAEs) were recorded and tabulated for comparison across SB-121 or placebo treatment, as were the vital signs and the hematology and blood chemistry parameters.

We conducted analysis of change from baseline in the Vineland-3 composite and domain scores, ABC subscale scores, and CGI-S utilizing a general linear model where the change score (i.e., the difference of post-28 days of treatment value from the pre-dosing value for each of the two periods) served as dependent variable. The difference in score was modeled as a function of treatment (SB-121 or placebo), study period (1 or 2), and the sequence of treatments (SB-121 in the first period or second). Subject was included in the model as a random effect and the sequence term measured potential crossover effect. If no crossover effect was noted for an outcome measure, then the adjusted (least squared) means for the treatments were given along with their difference and a p-value was assigned for the null hypothesis of no difference. Given the pilot nature of this analysis, p-values were not corrected for multiplicity. Given that the CGI-I is a Likert scale rating of improvement and is not administered at pre-treatment/baseline, CGI-I mean values post-treatment were compared between SB-121 and placebo To obtain eye tracking data, participants were seated in a quiet, dark room in front of a Tobii XL300 eye tracker at a distance of 60-65 cm from the eye tracker monitor. Each participant was presented with verbal instructions to look at the screen. The eye tracker was calibrated for each participant at the beginning of the session using the Tobii Studio “five-point calibration.” Successful calibration was ascertained via Tobii Studio’s automated validation procedure. A second attempt to calibrate was conducted if the participant did not successfully calibrate initially. Following calibration, participants completed a social interest paradigm, as previously published[39], where three silent 20 second side-by-side videos were presented with a social scene on one half of the screen and a geometric (i.e., non-social) pattern video on the other half. The side of the social scene video was pseudo-randomized and switched after each 20-second segment. Social scene preference ratio was calculated by dividing the time spent viewing the social scene videos by the total time spent viewing the social scene or geometric pattern videos. Thus, positive values indicate a “social preference” with more time spent looking at the social scene versus geometric pattern, whereas negative values indicate a “non-social preference” with more time looking at the geometric pattern.

Raw eye tracking data was exported from Tobii Studio and areas of interest (AOI) were created using MATLAB (version R2019a; The Mathworks, Inc., Natick, Massachusetts). The AOIs included the social scene or the geometric scene. The proportion of looking time was calculated by dividing the looking time to the AOI region by the total looking time to the geometric + social scenes. Proportion of valid looking was calculated by dividing the total looking time to anywhere on the screen divided by the total stimulus presentation time.

Participants were excluded if they had less than 35% valid looking data across the videos [39-41]. A generalized linear model was conducted with ratio of social versus non-social viewing as the dependent variable. The statistical analysis for all analyses, except eye tracking, were conducted using SAS® version 9.4 (SAS Institute Inc., Cary, NC). All eye tracking models were completed with SPSS. Cohen’s d effect sizes were included when appropriate.

## Results

Sixteen screening visits were conducted involving 15 individual participants. One participant screen failed due to concomitant proton pump inhibitor use and was subsequently rescreened and eventually randomized; see CONSORT Diagram in Figure 1 for study flow detail. Eight participants were randomized to receive placebo first and seven received SB-121 first. All randomized participants completed both treatment periods. Despite the protocol being open to male and female participants, all enrolled participants were male ranging in age from 15 to 27 years. Please see Table 1 for additional participant demographic details.

**Table 1:** Summary of Demographic and Baseline Characteristics (Intent to Treat Population). Abbreviations: ADOS-2 = Autism Diagnostic Observation Schedule, 2^nd^ edition; max = maximum; min = minimum; n = number of participants with data available; N = number of participants according to the first treatment sequence; SD = standard deviation; % = percentages were calculated based on N as the denominator. Note: Baseline was considered the last observation prior to dosing in Treatment Period 1. Drug abuse was considered positive if at least one of the parameters of drug abuse was positive and it was considered negative if all parameters of drug abuse were negative. ^a^ Body mass index (kg/m^2^) = body weight (kg)/height (m^2^)

| Characteristic<br>Statistic | SB-121 received first<br>(N = 7) | Placebo received first<br>(N = 8) | Total<br>(N = 15) |
| --- | --- | --- | --- |
| Age (years) |  |  |  |
| n | 7 | 8 | 15 |
| Mean (SD) | 20.1(1.46) | 19.9 (4.09) | 20.0 (3.05) |
| Median | 20.0 | 19.5 | 20.0 |
| Min, Max | 18, 22 | 15, 27 | 15, 27 |
| Sex, n (%) |  |  |  |
| Male | 7 (100.0) | 8 (100.0) | 15 (100.0) |
| Female | 0 | 0 | 0 |
| Race, n (%) |  |  |  |
| White | 7 (100.0) | 7 (87.5) | 14 (93.3) |
| Black or African American | 0 | 0 | 0 |
| Asian | 0 | 1 (12.5) | 1 (6.7) |
| Ethnicity, n (%) |  |  |  |
| Not Hispanic or Latino | 7 (100.0) | 8 (100.0) | 15 (100.0) |
| Height (cm) at Baseline |  |  |  |
| n | 7 | 8 | 15 |
| Mean (SD) | 177.96 (6.904) | 178.54 (6.280) | 178.27 (6.343) |
| Median | 179.40 | 180.75 | 179.70 |
| Min, Max | 169.0, 189.3 | 170.3, 186.2 | 169.0, 189.3 |
| Weight (kg) at Baseline |  |  |  |
| n | 7 | 8 | 15 |
| Mean (SD) | 69.67 (9.725) | 98.26 (40.006) | 84.92 (32.538) |
| Median | 69.00 | 91.15 | 75.80 |
| Min, Max | 59.0, 87.5 | 62.3, 188.9 | 59.0, 188.9 |
| Body Mass Index (kg/m<sup>2</sup>) at Baseline<sup>a</sup> |  |  |  |
| n | 7 | 8 | 15 |
| Mean (SD) | 22.11 (3.859) | 31.29 (14.552) | 27.01 (11.606) |
| Median | 21.20 | 29.25 | 21.60 |
| Min, Max | 19.0, 30.6 | 18.0, 65.1 | 18.0, 65.1 |

**Figure 1:**
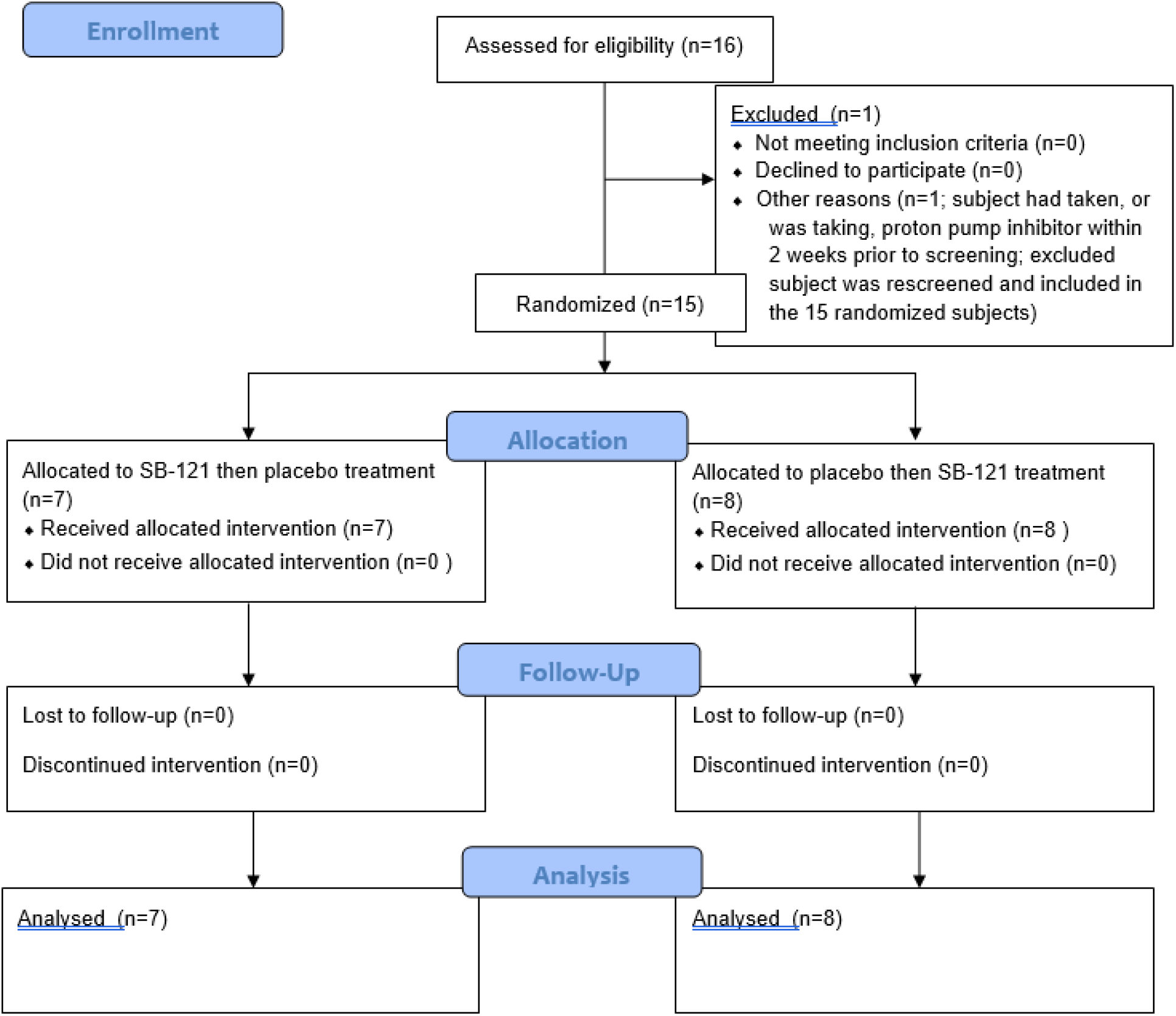
CONSORT Diagram.

Baseline WASI-II full scale values for the 15 randomized participants in the study showed a mean full-scale IQ of 88.66 (SD= 29.2; Range 40-128). The SCQ, used as an index of core autism symptom severity, had a mean score of 20.2 (SD= 8.06; Range 4-34). WASI and SCQ scores did not significantly differ for participants based on randomization order. Overall, the wide variance in WASI-II and SCQ scores in this study sample is consistent with that seen in patients with ASD clinically. Two participants scored ≤ 8 on the SCQ, but based on ADOS-2 and consensus diagnosis still met criteria for ASD. Regarding concomitant medication use, 15 (100.0%) received at least one concomitant medicine during the study (See Supplementary Information Tables S2 and S3 for full concomitant medication use data). The most frequently reported (≥10% of participants) concomitant medications for both the Treatment Period 1 and Treatment Period 2 included melatonin (4 [26.7%] participants); acamprosate, buspirone, clonidine, dexmethylphenidate hydrochloride, lisdexamphetamine mesilate, quetiapine, sertraline, metformin, vitamin D (3 [20.0%] participants each); amphetamine aspartate/amphetamine sulfate/dexamphetamine saccharate/dexamphetamine sulfate, guanfacine hydrochloride, methylphenidate hydrochloride, risperidone, vitamins, and fish oil (2 [13.3%] participants each).

Overall, use of SB-121 was well tolerated. Mean treatment compliance was similar between both treatment periods and treatment assignment. For SB-121 these were 92.2% and 90.4% for Periods 1 and 2 respectively; for placebo 95.7% and 84.3%. The treatment compliance data indicates that that the reconstitution and dosing instructions for the study drug (SB-121, placebo) were not a barrier to compliance and that it was well tolerated.

Treatment emergent adverse event (TEAE) rates were similar between SB-121 and placebo. During treatment period 1, among 11 participants with at least one TEAE, 5 received SB-121 (71.4%) and 6 (75.0%) received placebo. During treatment period 2, of 5 participants with at least one TEAE, 2 (25.0%) received SB-121 and 3 (42.9%) received placebo. Overall, during SB-121 treatment, 7 of 15 participants (46.7%) reported a total of 14 TEAEs, with the most common being gastrointestinal TEAEs reported by 3 (20.0%) participants. On the placebo arm, 9 (60.0%) of participants reported 23 TEAEs, with the most common being gastrointestinal TEAEs reported by 4 (26.7%) participants. Most of the adverse events reported on either arm were mild. There were no severe or serious adverse events during the trial. No participants discontinued study treatment due to adverse events. No participants had features of suspected bacteremia during the study. No changes from baseline in vital signs and safety laboratory and ECG parameters were noted with placebo or SB-121 treatment.

Regarding our analysis of outcome measures in evaluating potential clinical response to SB-121 treatment in these patients with autism, SB-121 treatment was associated with improvement in Vineland-3 scores compared to placebo in the entire subject sample (see Figure 2, Table 2). The Vineland-3 Adaptive Behavior Composite Score change from baseline (LS Means) in the SB-121 arm was statistically significant (p=0.03), and specifically within the Vineland-3 Daily Living Skills Domain (p=0.04). Additionally, there were small but positive SB-121 associated treatment effect size estimates of 0.32-0.41 across the Adaptive Behavior Composite Score, and the Socialization and Daily Living Skills domains.

**Table 2:** Summary of Vineland-3 Scores (change from baseline)

| Measure | Placebo |  |  | SB-121 |  |  | SB-121 minus Placebo |  |  | Approximate Treatment Effect Size |
| --- | --- | --- | --- | --- | --- | --- | --- | --- | --- | --- |
|  | LS Mean | SEM | P-value | LS Mean | SEM | P-value | LS Mean | SEM | P-value |  |
| Vineland Adaptive Behavior Composite Score | 1.05 | 2.06 | 0.61 | 4.87 | 2.06 | 0.03 | 3.81 | 3.21 | 0.26 | 0.32 |
| Vineland Socialization Domain | -0.14 | 2.79 | 0.96 | 5.57 | 2.79 | 0.06 | 5.71 | 4.81 | 0.26 | 0.32 |
| Vineland Daily Living Skills Domain | 0.53 | 2.42 | 0.83 | 5.37 | 2.42 | 0.04 | 4.84 | 3.14 | 0.15 | 0.41 |
| Vineland Communication Domain | 2.65 | 3.85 | 0.50 | 2.32 | 3.85 | 0.55 | -0.33 | 5.74 | 0.96 | -0.02 |

Abbreviations: LS = least squares; SEM = standard error of the mean.
For each measurement, the response was the difference between post-dose and pre-dose value. The adjusted, or LS mean for each treatment (SB-121 or placebo) and its standard error were derived from the model. The LS means differences between SB-121 and placebo (SB-121 LS mean minus Placebo LS mean) and their standard errors were also assessed. Statistical significance was set at a two-sided alpha = 0.05 and no adjustment was made for multiple comparisons. An approximate treatment effect was derived from the difference between the LS means for the SB-121 and placebo change from baseline and its standard error. All statistical analyses were conducted using SAS® statistical software version 9.4 for Windows (SAS Institute Inc., Cary, NC).

**Figure 2:**
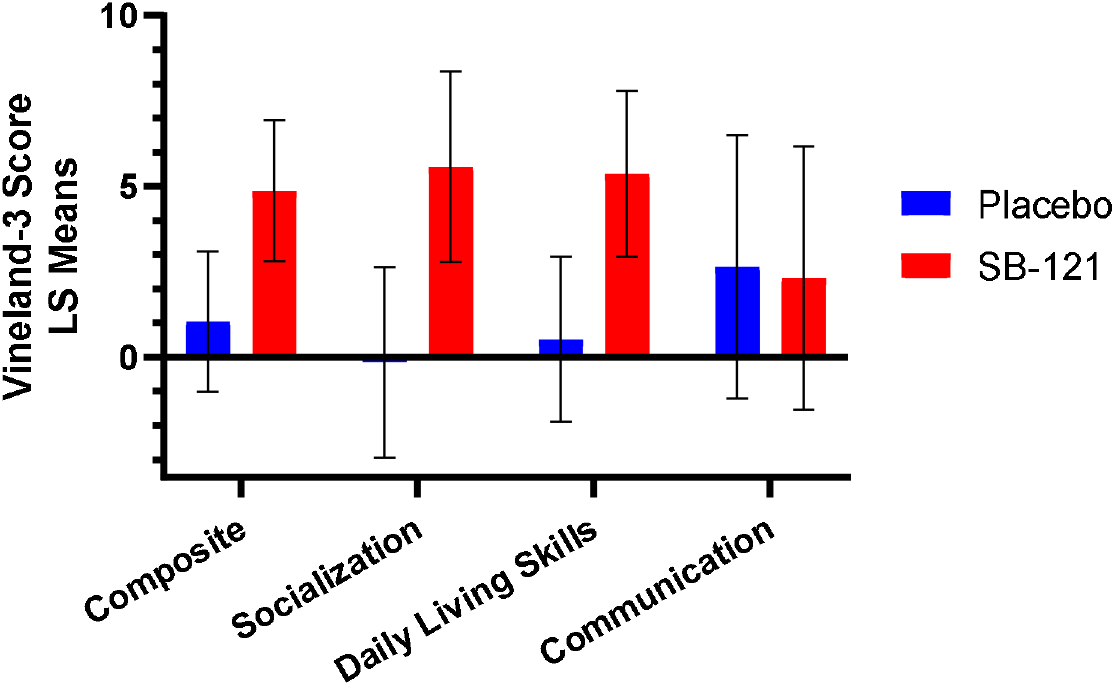
Vineland-3 Change from Baseline Scores (LS Means, SEM)

### Baseline Scores (LS Means, SEM)

Given the variable phenotypic presentation of our sample (based on WASI and SCQ), we examined individual-level responses and noted six participants had a robust Vineland-3 response indicated by an Adaptive Behavior Composite score change from baseline while on SB-121 versus placebo of ≥8 (Figure 3, Table 3, Supplementary Information Table S4). However, follow-up analyses comparing these so-called “robust responders” and “other subjects” on clinical measures or oxytocin levels at baseline did not reveal any significant differences. Considering improvement in this parameter by ≥8 compared to placebo as a robust response is a conservative estimate driven by prior clinical trial use of the Vineland in ASD where subject samples have noted Vineland Adaptive Behavior Composite score standard deviations ranging from ∼6 to ∼16 [42,43] indicating a ≥8 response would clinically represent in theory effect sizes from ∼0.5 to ∼1.25 in a responding population of subjects.

**Table 3:** Vineland-3 Robust Responders and Other Subjects (change from baseline).

|  | Robust Responders<br>n = 6 |  |  |  | Other Subjects<br>n = 9 |  |  |  |
| --- | --- | --- | --- | --- | --- | --- | --- | --- |
|  | Placebo |  | SB-121 |  | Placebo |  | SB-121 |  |
|  | Mean | SEM | Mean | SEM | Mean | SEM | Mean | SEM |
| Vineland Adaptive Behavior Composite Score | -0.33 | 4.97 | 10.50 | 4.06 | 1.33 | 2.18 | 1.89 | 2.60 |
| Vineland Socialization Domain | 0.50 | 5.86 | 15.83 | 4.29 | 4.56 | 5.64 | -6.11 | 3.27 |
| Vineland Daily Living Skills Domain | -1.33 | 2.73 | 14.50 | 1.91 | 3.00 | 2.25 | -1.44 | 1.40 |
| Vineland Communication Domain | -6.00 | 3.21 | 15.67 | 4.06 | 4.11 | 2.64 | -1.33 | 2.64 |

Abbreviations: SEM = standard error of the mean.

**Figure 3:**
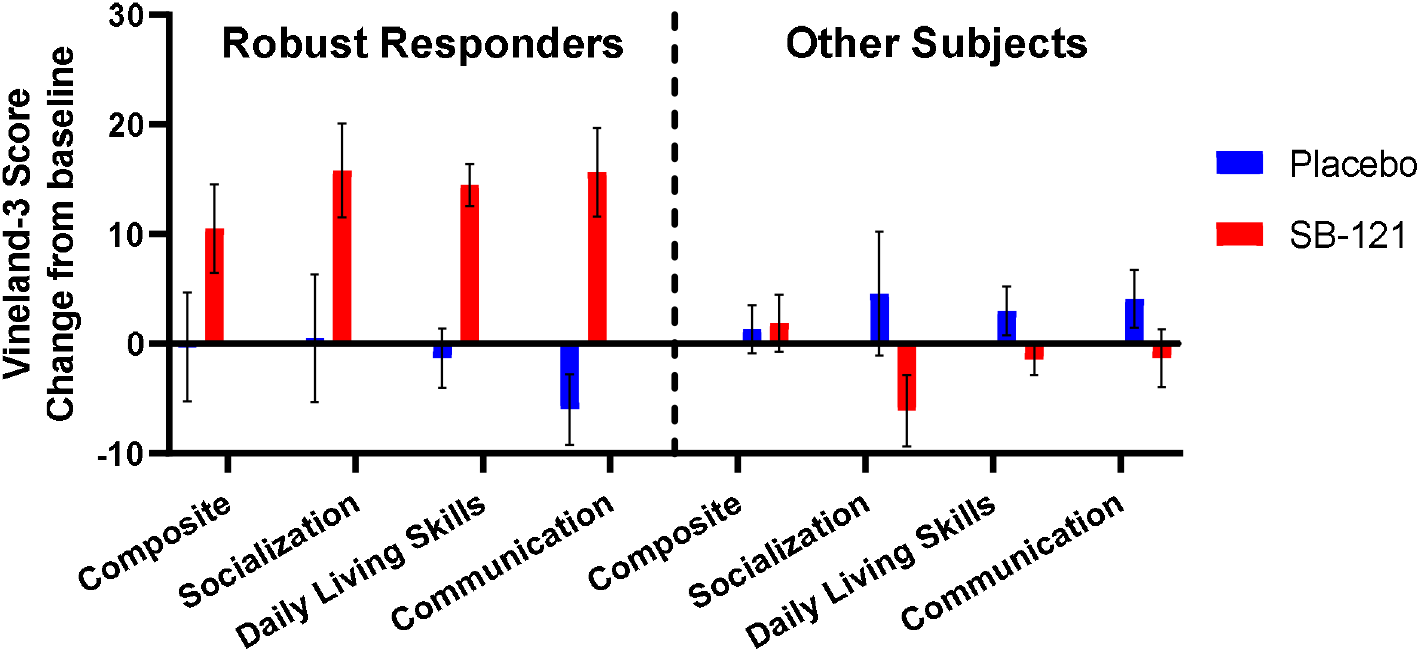
Vineland-3 Change from Baseline Scores - Robust Responders and Other Subjects (Mean, SEM)

Findings from the Aberrant Behavior Checklist (ABC) are presented in Table 4. The mean pre-treatment baseline raw scores across all ABC subscales were <10 across both treatment periods. Mean baseline scores for the Irritability and Stereotypic Behavior subscales were in the 3-4 range. These baseline findings could denote limited interfering behavioral challenges in this cohort of study participants. No SB-121-associated significant or directional changes were noted across all subscales of the ABC.

**Table 4:** Pre- and Post-Dose Data for ABC.

|  | Placebo |  |  |  | SB-121 |  |  |  |
| --- | --- | --- | --- | --- | --- | --- | --- | --- |
|  | Pre-Dose |  | Post-Dose |  | Pre-Dose |  | Post-Dose |  |
|  | Mean | SEM | Mean | SEM | Mean | SEM | Mean | SEM |
| Aberrant Behavior Checklist Irritability Subscale | 3.13 | 1.01 | 2.4 | 0.9 | 4 | 1.18 | 3.13 | 1.09 |
| Aberrant Behavior Checklist Social Withdrawal Subscale | 8.47 | 2.23 | 5.67 | 1.77 | 9.2 | 2.24 | 7.4 | 1.94 |
| Aberrant Behavior Checklist Stereotypic Behavior Subscale | 3.2 | 0.87 | 1.93 | 0.77 | 3.33 | 0.91 | 3.27 | 0.83 |
| Aberrant Behavior Checklist Hyperactivity Subscale | 8.07 | 1.89 | 6.07 | 1.52 | 7.40 | 1.57 | 7.67 | 1.54 |
| Aberrant Behavior Checklist Inappropriate Speech Subscale | 2.67 | 0.66 | 2.33 | 0.67 | 2.73 | 0.62 | 2.53 | 0.68 |

Abbreviations: ABC = Aberrant Behavior Checklist; SEM = standard error of the mean.

Regarding subscales of the CGI, with the CGI-S there were no relevant differences between placebo and SB-121 and the results suggested stable scores during SB-121 and placebo treatment. Mean CGI-I scores were also similar post-SB-121 versus placebo treatment (see Supplementary Information Table S5).

Utilizing eye tracking, there was a trend for increased social/geometric viewing ratio following SB-121 treatment compared to placebo (Figure 4), such that more time was spent looking at the social compared to geometric scene (p=0.1; SB-121 minus placebo of 107:1). This had a medium effect size of 0.61. When participants received SB-121 their ratio of viewing social versus non-social scenes was over 80:1, whereas viewing of non-social scenes versus social scenes was over 25:1 during placebo treatment. This suggests post-SB-121 treatment there was an increased social versus non-social preference during this task. Of note, there was a trending relationship between increased scores on the Vineland-3 Adaptive Behavior Composite and increased social scene viewing preference following SB-121 (Figure 5; r =.51, p =.09).

**Figure 4:**
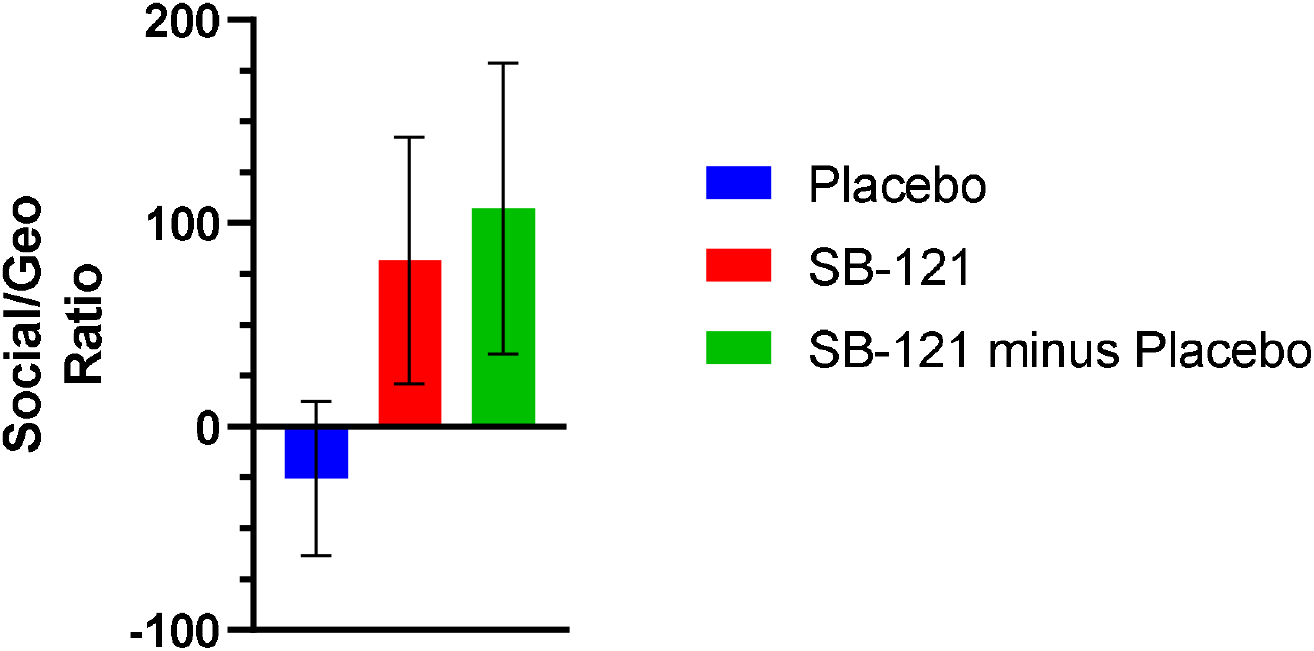
Eye Tracking Change from Baseline Score – Social/Geometric Ratio (LS Means, SEM)

**Figure 5:**
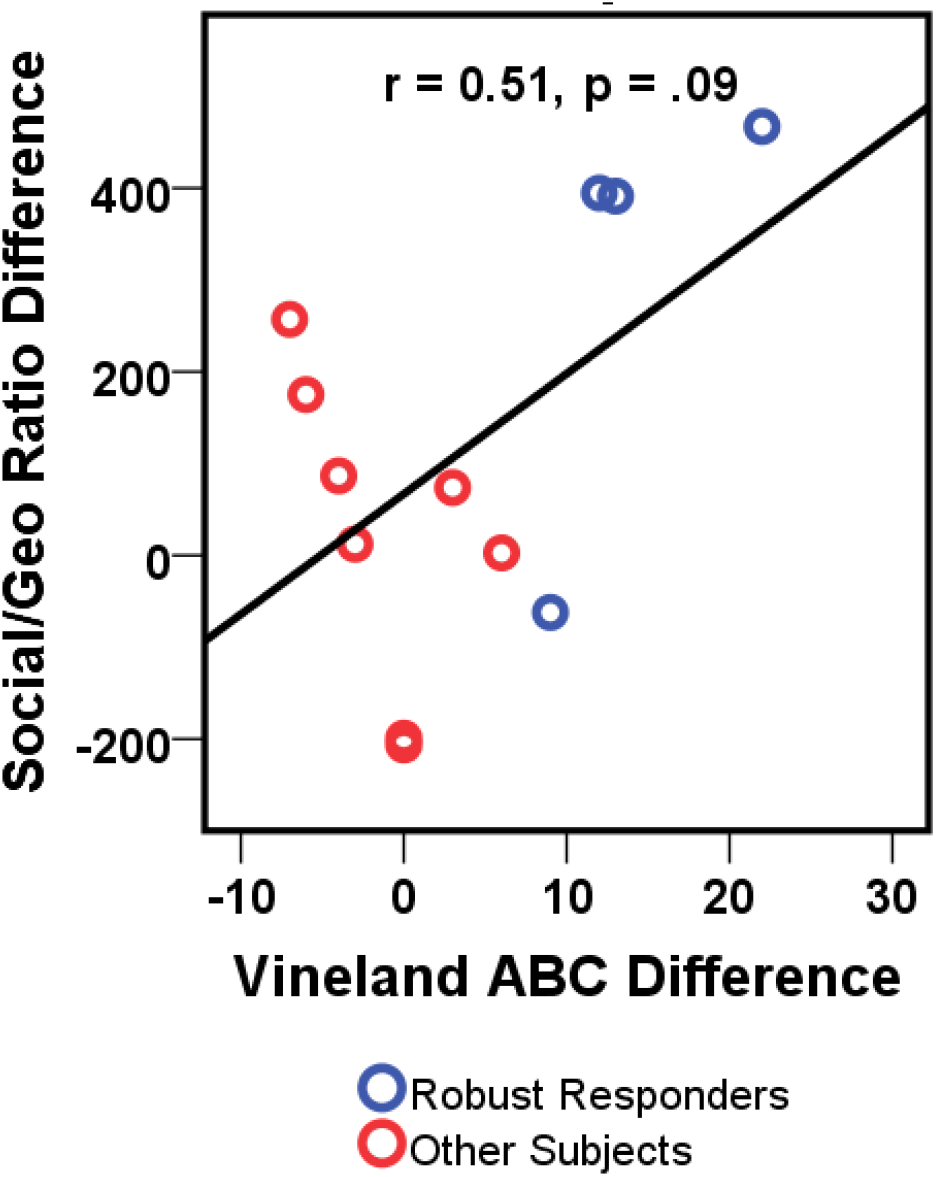
Relationship between Vineland-3 Change from Baseline and Social/Geometric Ratio Change from Baseline Following SB-121.

The mean (SD) percentage changes from baseline to Day 28 in plasma oxytocin levels for SB-121 and placebo groups were 111.63% (155.93) and 24.67% (80.94), respectively (Figure 6, Supplementary Information Table S6). The p-value for this difference is based on a paired t-test, p = 0.106, where the unpaired SB-121 is omitted from the calculation. However, percent change in oxytocin levels did not relate to changes in Vineland-3, ABC, or social viewing ratio (p’s>.05).

**Figure 6:**
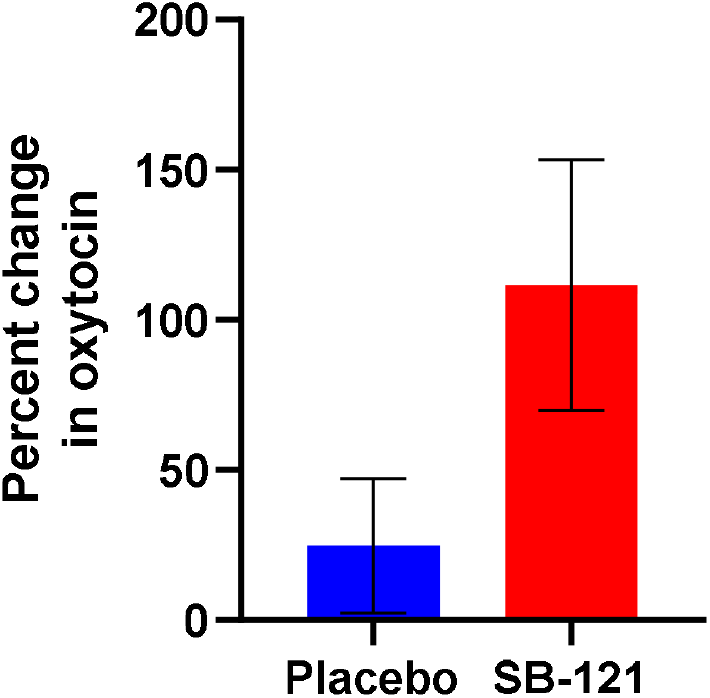
Plasma Oxytocin Percent Change from Baseline Values (Mean, SEM)

## Discussion

Overall SB-121 use was well tolerated and safe in 15 adolescents and adults with ASD. Treatment compliance in this pilot study was excellent which indicated that, given a standardized approach to reconstituting the formulation, patients would be able to take it as instructed. The enrolled participant sample was well representative of males with ASD broadly marked by variation in cognitive skills and severity of core ASD symptoms at baseline. Clinically, we documented clear directional improvements in adaptive behavior as measured by Vineland-3, which warrants replication in larger-scale study. Given the broad inclusion criteria for ASD utilized in this first-in-humans study, the Vineland-3 results are of particular interest as adaptive behavior deficits would be broadly expected in participants with ASD and this may have enabled this outcome measure to detect change in the context of significant baseline sample phenotypic heterogeneity.

The Vineland-3 findings are in contrast to behavioral findings from the ABC where no trends in SB-121-associated change were noted. This may be due to a lack of impact of SB-121 on interfering behaviors in ASD, but also may be due to significant likely floor effects with use of the ABC in this specific sample of autistic individuals who had very low ABC scores at baseline. Broad interpretation of ABC score change in ASD trials is difficult in cases where inclusion criteria do not prespecify ABC subscale or even an ABC total threshold score for study inclusion. It is possible that an early phase ASD clinical trial recruitment may bias towards enrollment of participants without significant interfering behavior that could preclude active study participation over a series of in person trial visits.

The finding that SB-121 was associated with a trend towards increased preference for viewing social versus non-social stimuli represents a quantitative, performance-based confirmation that this treatment could potentially enhance social interest in autistic individuals. Importantly, this was a trending quantitative correlate to improved adaptive behavior noted with treatment suggesting improved adaptive behavior as rated by a parent-rated measure may generalize to more real-world performance, especially within social interest. Thus, our findings warrant a larger-scale study for replication and extension.

Additional limitations of this report include a very small sample size that limited any ability to phenotypically define the subgroup of participants who appear to drive the overall positive directional findings of Vineland-3 ABC score and social viewing as measured by eye tracking. Further, since females were not represented in this study this omission is a study weakness and is likely a result of a small sample size and not stratifying enrollment by sex assigned at birth. Given the Phase I nature of the pilot study, had the sample included female participants, we would have not had any power to detect potential sex-associated differences in SB-121 tolerability or clinical response. Despite the gastrointestinal focus of the SB-121 intervention, we lack detailed assessment of GI symptoms in this trial, including potential quantitative change of stool features following SB-121 treatment. In future work it will be important to quantitatively evaluate GI symptoms at baseline and following treatment including potential use of quantitative evaluation of stool sampling given SB-121 direct exposure is limited to the gastrointestinal lumen. While a positive trend for plasma oxytocin measurements was observed when participants were taking SB-121, we note that there are significant limitations to plasma oxytocin measurements. Oxytocin is a notoriously difficult hormone to assay due to having a short half-life, being poorly immunogenic, and its tendency to bind to molecules in plasma[44].

Additionally, several participants had values below the lower limit of quantification at baseline, making it difficult to accurately quantify change with SB-121. Additionally, there are temporal differences between the release of central oxytocin and the accompanying secretion into the periphery; as such, plasma oxytocin levels should be treated with caution regarding its correlation with central oxytocin[45].

This Phase Ib study report on the first-in-humans use of SB-121 in male autistic participants provides data supporting future larger placebo-controlled studies of this compound in this autistic patients. The findings of a favorable safety profile of SB-121 in this sample of 15 individuals combined with significant improvements in Adaptive Behavior with treatment that is further corroborated by performance-based positive change in social engagement, indicates this treatment may be of benefit in ASD. It will be important in larger-scale studies to make efforts to rigorously phenotype the medical and gastrointestinal profile of participants in addition to classic clinical descriptions of behavior and cognition to potential define a subgroup who may best respond to treatment. This is of particular importance given the potential strong subgroup response we noted even in this small scale study that appeared to drive overall positive clinical change.

## Data Availability

All data produced in the present study are available upon reasonable request to the authors.

## Acknowledgements

We would like to acknowledge Gail E. Besner, M.D., Steven Goodman, PhD, Michael Bailey, PhD and Lauren Bakaletz, PhD from Nationwide Children’s Hospital, Columbus, OH as the inventors of the technology behind SB-121. We would also like to thank all participants, families and caregivers who contributed to this study.

## Author Contributions

LMS conducted aspects of study visits, participated in data analysis, manuscript writing and editing, and conceptualization of experiments. EGS, EVP, LB, and MW conducted aspects of study visits and participated in manuscript writing and editing. ML conducted aspects of study visits, participated in manuscript writing and editing, and participated in the conceptualization of experiments. PH is the study statistician and is responsible for the study analytic plan, participated in all study analyses, and participated in manuscript writing and editing. CAE conducted aspects of study visits, participated in manuscript writing and editing, and conceptualized elements of the experimental design employed in this manuscript. From Scioto Biosciences: SA conceptualized elements of the experimental design and formulated the protocol, reviewed the data and participated in data analyses, and contributed to the manuscript writing and review. JT, KM, MH, PH, and KW participated in data analysis, manuscript writing and editing.

## Data Availability Statement

Data is available on ClinicalTrials.gov (NCT049449) and can be requested from Scioto Biosciences.

## Additional Information

LMS, EGS, EVP, LB, MW, ML, PH and CAE are employees of Cincinnati Children’s Hospital Medical Center.

SA, JT, KM, MH, PH, and KW are employees and shareholders of Scioto Biosciences.

**Table S1:**
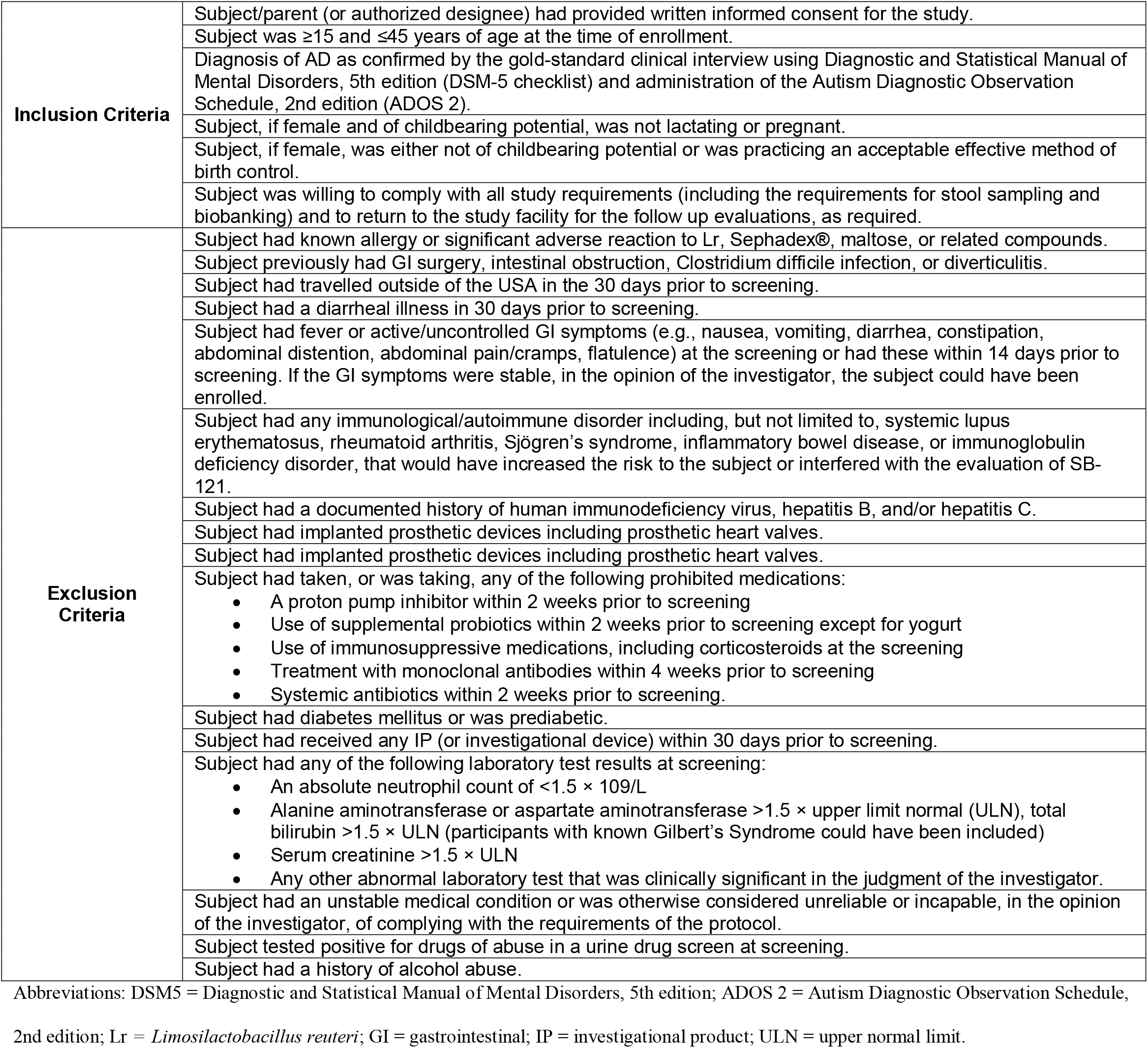
Inclusion and Exclusion Criteria.

**Table S2:**
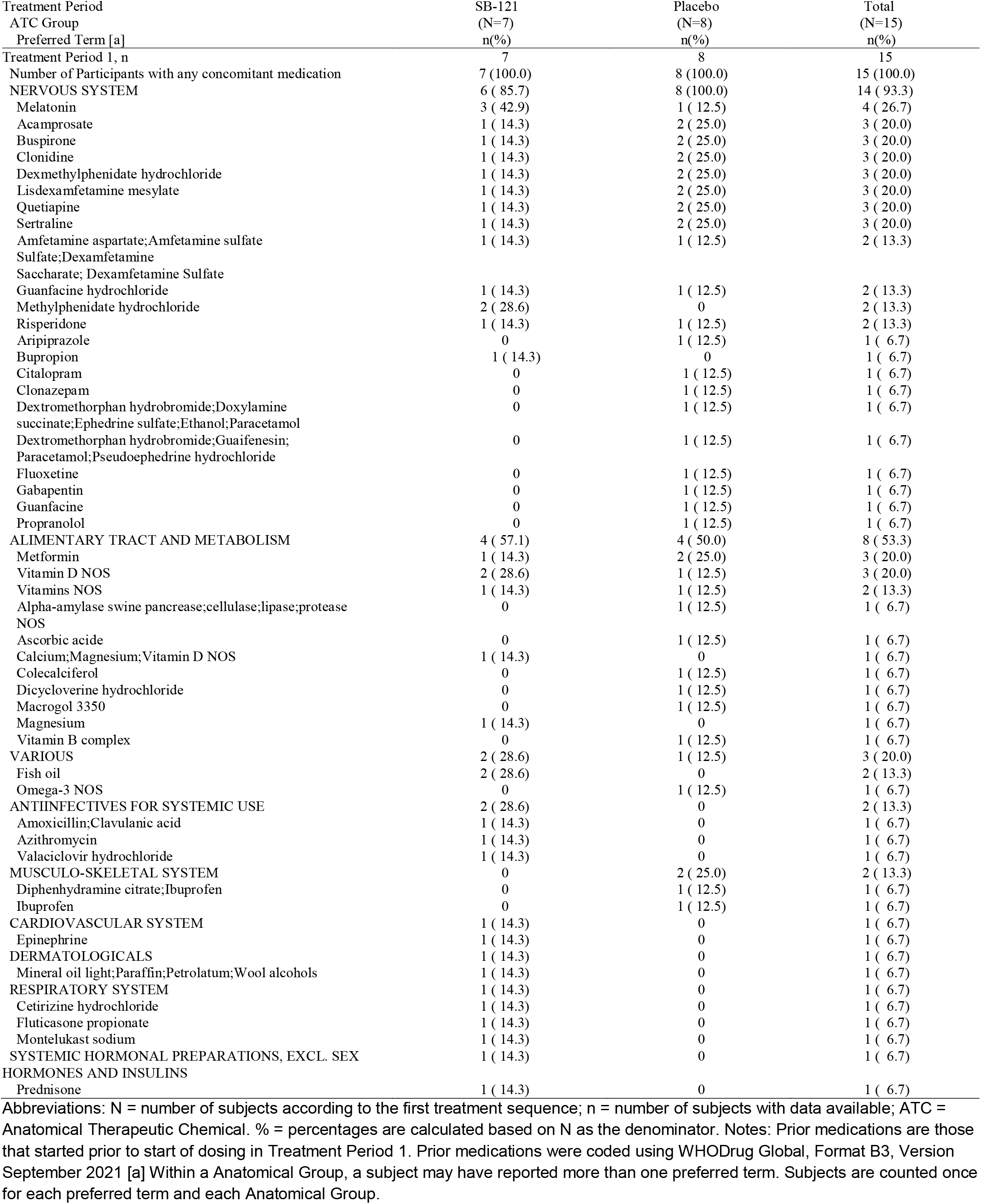
Summary of Concomitant Medications – Period 1.

| Treatment Period | SB-121 | Placebo | Total |
| --- | --- | --- | --- |
| ATC Group | (N=7) | (N=8) | (N=15) |
| Preferred Term [a] | n(%) | n(%) | n(%) |
| Treatment Period 1, n | 7 | 8 | 15 |
| Number of Participants with any concomitant medication | 7 (100.0) | 8 (100.0) | 15 (100.0) |
| **NERVOUS SYSTEM** | 6 ( 85.7) | 8 (100.0) | 14 ( 93.3) |
| Melatonin | 3 ( 42.9) | 1 ( 12.5) | 4 ( 26.7) |
| Acamprosate | 1 ( 14.3) | 2 ( 25.0) | 3 ( 20.0) |
| Buspirone | 1 ( 14.3) | 2 ( 25.0) | 3 ( 20.0) |
| Clonidine | 1 ( 14.3) | 2 ( 25.0) | 3 ( 20.0) |
| Dexmethylphenidate hydrochloride | 1 ( 14.3) | 2 ( 25.0) | 3 ( 20.0) |
| Lisdexamfetamine mesylate | 1 ( 14.3) | 2 ( 25.0) | 3 ( 20.0) |
| Quetiapine | 1 ( 14.3) | 2 ( 25.0) | 3 ( 20.0) |
| Sertraline | 1 ( 14.3) | 2 ( 25.0) | 3 ( 20.0) |
| Amphetamine aspartate;Amphetamine sulfate | 1 ( 14.3) | 1 ( 12.5) | 2 ( 13.3) |
| Sulfate;Dexamfetamine |  |  |  |
| Saccharate; Dexamfetamine Sulfate |  |  |  |
| Guanfacine hydrochloride | 1 ( 14.3) | 1 ( 12.5) | 2 ( 13.3) |
| Methylphenidate hydrochloride | 2 ( 28.6) | 0 | 2 ( 13.3) |
| Risperidone | 1 ( 14.3) | 1 ( 12.5) | 2 ( 13.3) |
| Aripiprazole | 0 | 1 ( 12.5) | 1 ( 6.7) |
| Bupropion | 1 ( 14.3) | 0 | 1 ( 6.7) |
| Citalopram | 0 | 1 ( 12.5) | 1 ( 6.7) |
| Clonazepam | 0 | 1 ( 12.5) | 1 ( 6.7) |
| Dextromethorphan hydrobromide;Doxylamine succinate;Ephedrine sulfate;Ethanol;Paracetamol | 0 | 1 ( 12.5) | 1 ( 6.7) |
| Dextromethorphan hydrobromide;Guaifenesin;Paracetamol;Pseudoephedrine hydrochloride | 0 | 1 ( 12.5) | 1 ( 6.7) |
| Fluoxetine | 0 | 1 ( 12.5) | 1 ( 6.7) |
| Gabapentin | 0 | 1 ( 12.5) | 1 ( 6.7) |
| Guanfacine | 0 | 1 ( 12.5) | 1 ( 6.7) |
| Propranolol | 0 | 1 ( 12.5) | 1 ( 6.7) |
| **ALIMENTARY TRACT AND METABOLISM** | 4 ( 57.1) | 4 ( 50.0) | 8 ( 53.3) |
| Metformin | 1 ( 14.3) | 2 ( 25.0) | 3 ( 20.0) |
| Vitamin D NOS | 2 ( 28.6) | 1 ( 12.5) | 3 ( 20.0) |
| Vitamins NOS | 1 ( 14.3) | 1 ( 12.5) | 2 ( 13.3) |
| Alpha-amylase swine pancrease;cellulase;lipase;protease NOS | 0 | 1 ( 12.5) | 1 ( 6.7) |
| Ascorbic acid | 0 | 1 ( 12.5) | 1 ( 6.7) |
| Calcium;Magnesium;Vitamin D NOS | 1 ( 14.3) | 0 | 1 ( 6.7) |
| Colecalciferol | 0 | 1 ( 12.5) | 1 ( 6.7) |
| Dicycloverine hydrochloride | 0 | 1 ( 12.5) | 1 ( 6.7) |
| Macrogol 3350 | 0 | 1 ( 12.5) | 1 ( 6.7) |
| Magnesium | 1 ( 14.3) | 0 | 1 ( 6.7) |
| Vitamin B complex | 0 | 1 ( 12.5) | 1 ( 6.7) |
| **VARIOUS** | 2 ( 28.6) | 1 ( 12.5) | 3 ( 20.0) |
| Fish oil | 2 ( 28.6) | 0 | 2 ( 13.3) |
| Omega-3 NOS | 0 | 1 ( 12.5) | 1 ( 6.7) |
| **ANTIINFECTIVES FOR SYSTEMIC USE** | 2 ( 28.6) | 0 | 2 ( 13.3) |
| Amoxicillin;Clavulanic acid | 1 ( 14.3) | 0 | 1 ( 6.7) |
| Azithromycin | 1 ( 14.3) | 0 | 1 ( 6.7) |
| Valaciclovir hydrochloride | 1 ( 14.3) | 0 | 1 ( 6.7) |
| **MUSCULO-SKELETAL SYSTEM** | 0 | 2 ( 25.0) | 2 ( 13.3) |
| Diphenhydramine citrate;Ibuprofen | 0 | 1 ( 12.5) | 1 ( 6.7) |
| Ibuprofen | 0 | 1 ( 12.5) | 1 ( 6.7) |
| **CARDIOVASCULAR SYSTEM** | 1 ( 14.3) | 0 | 1 ( 6.7) |
| Epinephrine | 1 ( 14.3) | 0 | 1 ( 6.7) |
| **DERMATOLOGICALS** | 1 ( 14.3) | 0 | 1 ( 6.7) |
| Mineral oil light;Paraffin;Petrolatum;Wool alcohols | 1 ( 14.3) | 0 | 1 ( 6.7) |
| **RESPIRATORY SYSTEM** | 1 ( 14.3) | 0 | 1 ( 6.7) |
| Cetirizine hydrochloride | 1 ( 14.3) | 0 | 1 ( 6.7) |
| Fluticasone propionate | 1 ( 14.3) | 0 | 1 ( 6.7) |
| Montelukast sodium | 1 ( 14.3) | 0 | 1 ( 6.7) |
| **SYSTEMIC HORMONAL PREPARATIONS, EXCL. SEX HORMONES AND INSULINS** | 1 ( 14.3) | 0 | 1 ( 6.7) |
| Prednisone | 1 ( 14.3) | 0 | 1 ( 6.7) |

Abbreviations: N = number of subjects according to the first treatment sequence; n = number of subjects with data available; ATC = Anatomical Therapeutic Chemical. % = percentages are calculated based on N as the denominator. Notes: Prior medications are those that started prior to start of dosing in Treatment Period 1. Prior medications were coded using WHODrug Global, Format B3, Version September 2021 [a] Within a Anatomical Group, a subject may have reported more than one preferred term. Subjects are counted once for each preferred term and each Anatomical Group.

**Table S3:**
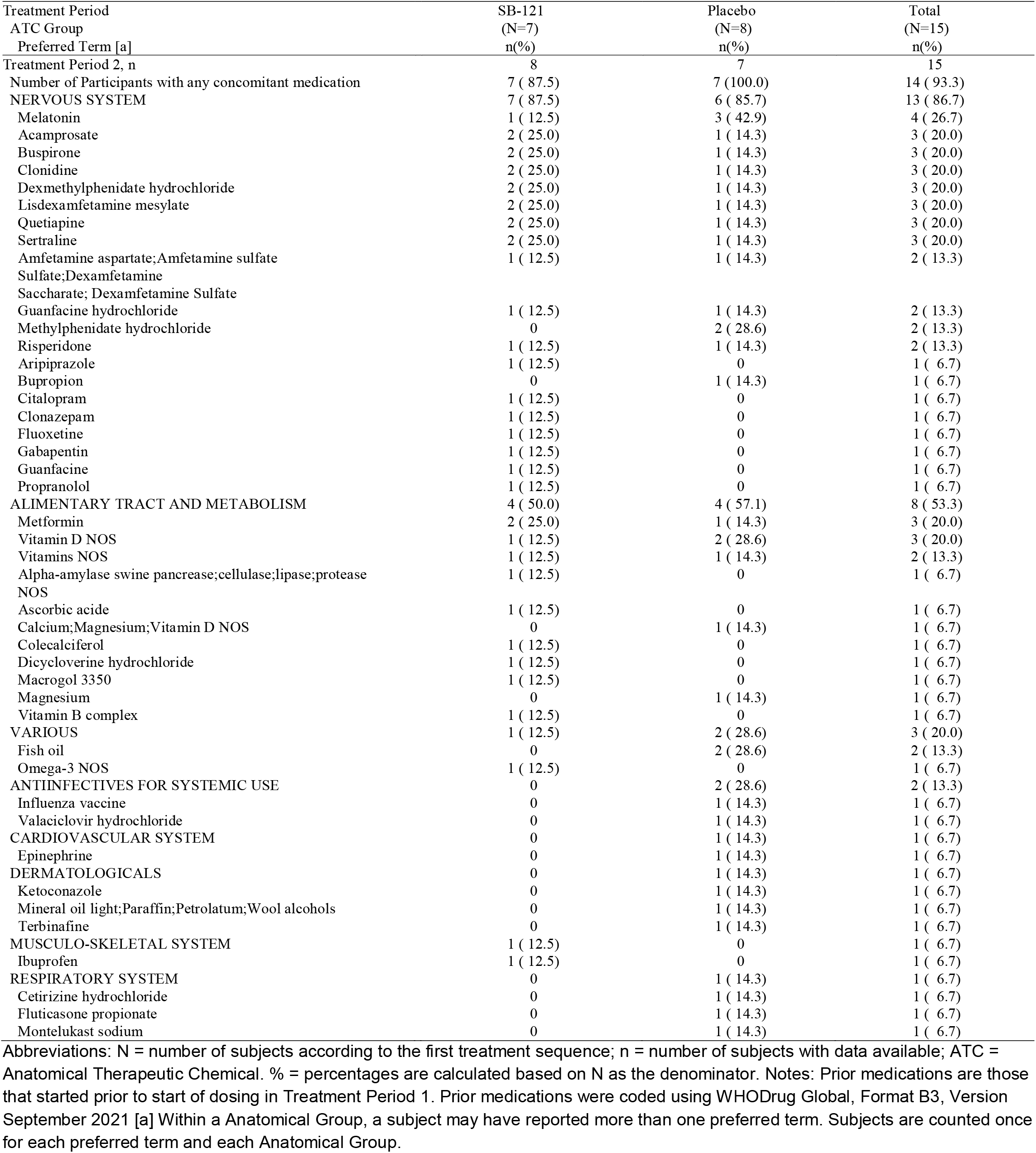
Summary of Concomitant Medications – Period 2.

**Table S4:** Individual Vineland Adaptive Behavior Scale Scores (change from baseline).

| Subject ID | Vineland Adaptive Behavior Composite Score |  |  | Vineland Socialization Domain |  |  | Vineland Daily Living Skills Domain |  |  | Vineland Communication Domain |  |  |
| --- | --- | --- | --- | --- | --- | --- | --- | --- | --- | --- | --- | --- |
| | SB-121 | Placebo | $\Delta^a$ | SB-121 | Placebo | $\Delta^b$ | SB-121 | Placebo | $\Delta^b$ | SB-121 | Placebo | $\Delta^b$ |
| 001 | 12 | -12 | 24 | 24 | -21 | 45 | 8 | 0 | 8 | 4 | -12 | 16 |
| 002 | 13 | 4 | 9 | 9 | -2 | 11 | 8 | 20 | -12 | 24 | -8 | 32 |
| 003 | -4 | 11 | -15 | 4 | 9 | -5 | -8 | 0 | -8 | -8 | 28 | -36 |
| 004 | 13 | -6 | 19 | 23 | -5 | 28 | 17 | -11 | 28 | 4 | -6 | 10 |
| 005 | 6 | 2 | 4 | 3 | -8 | 11 | 18 | 11 | 7 | -3 | 4 | -7 |
| 006 | 0 | 3 | -3 | -14 | 5 | -19 | 4 | -5 | 9 | 10 | 7 | 3 |
| 008 | 9 | -1 | 10 | 10 | 2 | 8 | 3 | -4 | 7 | 15 | -1 | 16 |
| 009 | 22 | 1 | 21 | 2 | -6 | 8 | 27 | 6 | 21 | 30 | 2 | 28 |
| 010 | -3 | -8 | 5 | -1 | 0 | -1 | 3 | -6 | 9 | -13 | -21 | 8 |
| 011 | 3 | 8 | -5 | 3 | 18 | -15 | 3 | 6 | -3 | 2 | 0 | 2 |
| 012 | 18 | 6 | 12 | 26 | -4 | 30 | 0 | -13 | 13 | 18 | 28 | -10 |
| 013 | -6 | -5 | -1 | -6 | 6 | -12 | 2 | -6 | 8 | -14 | -16 | 2 |
| 014 | -2 | 0 | -2 | -13 | 5 | -18 | 5 | -2 | 7 | 0 | -2 | 2 |
| 015 | -7 | 4 | -11 | 4 | -6 | 10 | -8 | 6 | -14 | -23 | 14 | -37 |
| 016 | 0 | 12 | -12 | 8 | 8 | 0 | -2 | 8 | -10 | -6 | 27 | -33 |

<sup>a</sup> Participants 001, 002, 004, 008, 009 and 012 were the 6 participants with a placebo adjusted change from baseline of $\geq 8$ on SB-121 on the Adaptive Behavior Composite score.
<sup>b</sup> SB-121 minus Placebo.

**Table S5:** Pre- and Post-Dose Data for CGI Scales.

|  | Placebo |  |  |  | SB-121 |  |  |  |
| --- | --- | --- | --- | --- | --- | --- | --- | --- |
|  | Pre-Dose |  | Post-Dose |  | Pre-Dose |  | Post-Dose |  |
|  | Mean | SEM | Mean | SEM | Mean | SEM | Mean | SEM |
| Clinical Global Impressions Severity Subscale | 3.87 | 0.13 | 3.93 | 0.12 | 3.93 | 0.12 | 3.87 | 0.13 |
| Clinical Global Impressions Improvement Subscale | NA | NA | 3.6 | 0.16 | NA | NA | 3.47 | 0.26 |

**Table S6:** Summary of Overall Plasma Oxytocin Levels.

|  | SB-121 |  |  | Placebo |  |  |
| --- | --- | --- | --- | --- | --- | --- |
|  | Value at Visit | Change from Baseline | Percentage Change from Baseline | Value at Visit | Change from Baseline | Percentage Change from Baseline |
| **Oxytocin (pg/mL)** |  |  |  |  |  |  |
| Baseline |  |  |  |  |  |  |
| n | 15 |  |  | 15 |  |  |
| Mean (SD) | 37.0647<br>(23.54037) |  |  | 41.2383<br>(22.70178) |  |  |
| Median | 27.0600 |  |  | 34.5700 |  |  |
| Min, Max | 15.625,<br>74.690 |  |  | 15.625,<br>101.880 |  |  |
| End of Treatment |  |  |  |  |  |  |
| n | 14 | 14 | 14 | 13 | 13 | 13 |
| Mean (SD) | 53.3025<br>(23.05427) | 17.9539<br>(30.42421) | 111.6278<br>(155.93306) | 44.2181<br>(18.15442) | 0.8515<br>(22.75805) | 24.6667<br>(80.94121) |
| Median | 56.6650 | 17.4525 | 55.5703 | 45.9900 | 1.6500 | 3.6748 |
| Min, Max | 15.625,<br>103.930 | -31.290,<br>84.860 | -42.258,<br>444.992 | 15.625,<br>71.260 | -43.660,<br>37.390 | -44.142,<br>191.842 |

Abbreviations; n = number of participants with data available; SD = Standard deviation; Min = Minimum; Max = Maximum; LLQ = Lower limit quantification.

